# Exploring facilitators for a transition from alternative and complementary therapies to evidence-based treatments in Ugandan first-episode psychosis patients

**DOI:** 10.1101/2022.02.21.22270378

**Authors:** Emmanuel K. Mwesiga, Andrew S. Ssemata, Ann Jacquelline Nakitende, Linnet Ongeri, Aggrey Semeere, Rachel Loewy, Susan Meffert, Noeline Nakasujja

## Abstract

**Introduction:** Most patients with psychotic disorders in Africa initially use alternative and complementary therapies leading to delays accessing evidence-based treatments. This delay, known as the duration of untreated psychosis (DUP), is associated with suboptimal response and reduced efficacy in short-term and long-term outcomes. In this study, we explore facilitators for transitioning from alternative and complementary therapies to evidence-based treatments for psychotic disorders, comparing these facilitators between patients and their caregivers.

**Methods:** The study was conducted at Butabika hospital in Kampala, Uganda. Participants included patients with first-episode psychosis who had used alternative and complementary therapies and their caregivers. An exploratory qualitative design using in-depth interviews was performed with thematic analysis used to analyze the data.

**Results:** We interviewed eight patients and eight caregivers. The key themes that led to a switch were the high cost of alternative therapies, a desire to know the cause of the illness, seeking better care and the influence of friends and community members. Themes were similar among the participants and their caregivers except for stigma, which was only found among the caregivers.

**Conclusions:** The reasons for transitioning from alternative and complementary therapies to evidence-based treatments are similar between patients and caregivers, except for stigma that is a more important factor for transition among caregivers. Since this was an exploratory study, additional research with a larger sample and more diverse population would enable a deeper understanding of these factors and guide the development of interventions to reduce DUP and improve outcomes for psychotic disorders.

## Introduction

More than half of the patients with a psychotic disorder in Africa initially use alternative and complementary therapy (ACT) as their first treatment before switching to evidence-based treatments (1-3). A more significant proportion of patients with psychotic disorders use ACT than other disorders like HIV/AIDS (4). ACT involves the use of non-conventional approaches as a means of treatment (5). For psychotic disorders, these may include religious healers, religious rituals, herbalists and traditional healers (2, 6, 7). Preference for alternative therapies is premised on spiritual attributions as the cause of a psychotic illness (8). This differs from evidence-based treatments (EBTs) like antipsychotic medication, psychological and social interventions premised on a biopsychosocial model of illness (9-13).

The use of alternative and complementary therapies and delays in initiation of evidence-based treatments like antipsychotic medication is associated with prolonged duration of untreated psychosis (DUP) (8, 14). In Sub-Saharan Africa (SSA), psychosis patients have a mean DUP of up to five years, compared to one year in high-income countries (HICs) (15, 16). Shorter DUP like that seen in HICs is associated with lower symptom severity, higher cognitive functioning, decreased substance use and less disability (15-17). Furthermore, the WHO has recommended a target DUP of 90 days(18). The first step toward reducing DUP in this population is understanding why patients with psychosis and their families switch to evidence-based treatments after initially using ACT.

Most literature on alternative and complementary therapy use in patients with psychotic disorders has not explored the reasons for transitioning to evidence-based treatments. Approximately 35% of patients using ACT transition to EBTs (19). These patients’ initial presentation to hospitals for EBTs is characterized by severe illness presentations (1). However, the reasons for the transition from ACT to EBTs are unclear (1, 19, 20). It is also unclear if the reasons for the transition are similar between patients and caregivers. This knowledge is essential for services providing evidence-based treatments to improve future engagements (21).

This study aimed to explore the reasons for the transition from alternative and complementary therapy to evidence-based treatments in patients with psychotic disorders from Uganda. Specifically, we wanted to understand the reasons for the switch; and if these reasons differ among patients and caregivers.

## Methods

### Setting

The study was conducted at Butabika National Psychiatric Referral and Teaching hospital of Uganda (BNPRTH). The setting has been described previously (22). Briefly, it is a 200-bed hospital that serves as the leading tertiary psychiatric institution. The selection of BNPRTH was informed by the desire to access individuals from different backgrounds. It has a wide catchment area of patients from all regions in the country, which improved the applicability of study findings across various backgrounds.

### Study design

This was an exploratory qualitative study design that used in-depth interviews (IDIs) guided by a semi-structured interview guide to explore motivations for transitioning from ACT to evidence-based treatments in patients with psychotic disorders. IDIs were considered appropriate for generating data on personal, lived experiences among the caregivers and patients and thematic analysis.

### Sample

The participants were already enrolled in a study on cognition at the first episode of psychosis (FEP) (19). The participants in the previous study had provided informed consent and permission to be contacted for future studies. They were invited to participate in the study if they could speak English or Luganda, had used ACT before attending the hospital and were able to come along with a caregiver. The caregiver was someone who had actively been involved in the care of the patient. Individuals were purposively selected and recruited until a point of saturation (no new information emerged from the collected data) was reached.

### Data collection

The study was conducted from a quiet and conducive facility at BNPRTH between April and September 2019 among 16 individuals (eight patients and eight caregivers). The research nurse invited potentially eligible patients and caregivers to participate in the in-depth interviews conducted by two authors (ASS and AJN). The interviewers obtained informed consent and conducted in-depth interviews for each patient and their caregiver separately. In-depth interviews were conducted in English or Luganda (a local dialect), using a semi-structured interview guide with open-ended questions and probes lasting 45-70 minutes. The interviews were audio-recorded with consent from the participants and later transcribed verbatim alongside detailed hand-written notes by the interviewer to supplement the audio data. Three in-depth interviews (patients n=2, caregiver n=1) conducted in Luganda were translated into English.

### Data analysis

We employed thematic analysis and developed a matrix scheme to interpret patterns of meaning from a rich and detailed yet complex account of data corpus as advanced by Braun and Clarke (2006) (23). The six steps followed included familiarization, coding, generating themes, reviewing themes, defining and naming themes, and writing up. Interview transcripts were reviewed alongside the audio recordings and interview notes to check for accuracy and enable familiarization of the data by the two authors (ASS, AJN). The first author (EKM) was an independent reviewer as he was not involved in the data collection process. The initial two transcripts from each participant category were read several times by two authors (ASS and AJN) independently through a process of open coding until no new issues were identified. After reaching an agreement on the selected codes, these were organized into categories and sub-categories to develop a codebook with main and sub-themes. Any new ideas identified from subsequent transcripts were coded and included in the codebook and discrepancies were discussed and resolved by involving the third reviewer (EKM). The codebook was reviewed by the first author (EKM) to ensure completeness of data retrieval, then later used to analyze the remaining transcripts and the codebook by two other authors (ASS and AJN). The data collection process continued upon completion of coding and analysis of the previously collected interviews to ensure we collected sufficient information to address the research question until saturation. Saturation was attained when most of the themes emerged, and no new information arose (24). Relevant excerpts were identified and are reported in the findings section to illustrate and represent the participants’ views.

## Findings

We obtained data saturation after interviewing 16 participants (8 patients and 8 caregivers). The median age of the patients was 28 years (IQR 22-34), while the median age of the caregivers was 39.5years (IQR 31-45.5). The patients comprised four females and four males, while the caregivers were three females and five males. Caregivers included two spouses, two parents, one sibling, one close friend and two aunt/uncles. One caregiver had previously suffered from a psychotic disorder, and 6/8 caregivers were currently employed. The themes from the participant interviews are summarized in Table 1.

**Table 1:** Summary of themes and sub-themes observed in the interviews.

|  | Theme | Sub-themes |
| --- | --- | --- |
| 1. | **Dissatisfaction with ACT** | *a) Significant and worsening mental symptoms* |
|  |  | *b) Ineffectiveness of the alternative treatment* |
|  |  | *c) Multiple demands and cost of alternative and complementary therapy* |
| 2. | **Desire to know the cause of the illness and seek better care** | *a) Desperation in seeking the cause of illness* |
|  |  | *b) Desire for a cure and better medical treatment.* |
|  |  | *c) Knowledge of where to seek evidence-based treatments* |
| 3. | **Influence of friends/community members** | *a) Exposure to lived positive experience of Western medical treatment from Butabika* |
|  |  | *b) Receiving recommendations and referral* |
|  |  | *c) Avoiding stigma from the community* |

### 1. DISSATISFACTION WITH ACT

#### a. Multiple demands and cost of ACT

The notion ‘no money, no care’ resonated strongly with majority of the participants and caregivers who sought ACT care before switching to evidence-based treatments.

> *“When they used to take me there [mentions location of a traditional healer], they used to ask us a lot of things like goats, chicken, money, clothes and they could tell us that I was attacked by a small goddess called Nakayima who needed to dress up in a gomesi [female traditional attire] every time we went there. It was so expensive as they demanded for two gomesi and things like chicken. After a while I realized that our money was being wasted and you could not get any service without meeting the demands… The good thing here in Butabika when medicines are available, it is free of charge.”* ***female participant***.
>
> *“When you look at the [mentions religious place] nowadays there are very many false service providers, someone will come out and say that he is going to help you but you first have to pay a large amount of money. They don’t know how the sickness came about; the effects and then they say you are going to be well meanwhile they have ‘eaten’ [taken] my money. Time reached when I didn’t have money and he was not improving so I decided to come to the professional people”*. ***male caregiver***.

#### b. Ineffectiveness of ACT

Participants decided to try out evidence-based treatments on finding ACT was ineffective and failed to resolve the condition.

> *“The challenge we faced that made us come here for care was that we were given some things [herbs and charms from a traditional healer] to help as you know our things [using African traditional healers] but the condition failed to improve and she actually kept getting worse. We decided those things are not working, let’s try the medical doctors.”* ***female caregiver***.
>
> *“They took me to some witches, and they told me to take some herbs for some time. They were forcing me to take herbs all the time, but I never saw any improvement. So, when they [caregivers] saw that all was not working they just ended bringing me here to Butabika after spending lots of money.”* ***male participant***

#### c. Delayed identification as mental illness

Initially participants had not classified their symptoms as mental illnesses and only resorted to evidence-based treatments when symptoms did not improve.

> *“Before it had started, we did not come here we went to only church. The next option was to bring him to Butabika that is where they rehabilitate mental disorders, and whatever will come from Butabika it’s what we shall go with”* ***Female caregiver***.
>
> *“Ok, the change they saw in me made them think I was ‘mental’. They started thinking that I was going to get mad because I had totally changed, and I would not sleep. I lost appetite, and I would not eat, but I would just be there…I would drink sodas, Splash [packed fruit juice], water, but I would not eat. So those are some of the things that prompted them to think that I had got a mental disorder because they used to see that I was disorganizing the property at home so they decided to bring me here [Butabika hospital]. Actually, that night before bringing me here my relative wanted to get a rope and tie me up.”* ***female participant***.

### 2. DESIRE TO KNOW THE CAUSE OF THE ILLNESS AND SEEK BETTER CARE

Participants expressed a desire to know the cause of illness and better ways to manage the symptoms.

#### a. Desperation in seeking the cause of illness

Participants also noted that they were desperate to seek answers, especially to understand the cause of the patient’s illness.

“*I think after trying all these things, they were tired and needed a solution, so this time they took me to the hospital and I was given some treatment. They wanted to find out what is exactly was disturbing me. We all could not understand it and no one where we had been could explain. So they brought me here.”* ***female participant***.

“*But what forced me to bring my child here is I really wanted to see that if his head is normal or there’s something that is not right on his head. When he had that problem before and he was prayed for, the situation normalized. This time I told them [the people who were praying] lets first take him to hospital and examine him to see what disturbs this child. If the situation fails, there also will give us a way forward”*. ***female caregiver***

#### b. Desire for a cure and better medical treatment

Both patients and caregivers reported that the desire for better and quicker treatment to resolve the mental condition was another reason for transitioning to evidence-based treatments.

> *“I was eventually brought here because I wanted to be healthy with nothing disturbing my head. The drugs here treat the mental problem, and I think when I stop taking them, I will experience a mental disorder… This medicine is to refresh the brain and also to cure the wounds inside.”* ***male participant***.
>
> *“Personally, I believe that I can pray alone, and God listens to my prayers. But I brought her to the hospital because this is the most skilled hospital in Uganda while at the same time I prayed so that we were not praying only so that God can use the doctors to treat her, maybe get a cure and also make miracles happen again just like those who use herbs and African magic [witchcraft]”* ***male caregiver***

### 3. INFLUENCE OF FRIENDS/COMMUNITY MEMBERS

Family or friends were essential in ensuring that patients and their caregivers sought care and influenced the transition to evidence-based treatments in various ways.

#### a. Lived positive experience of Western medical treatment from a mental health facility

Previous patients and caregivers with lived experiences about using evidence-based treatment and services at the National Mental Health referral hospital were crucial in convincing patients to switch to evidence-based treatments.

> *“At the church where they used to take me for prayers, there was a pastor who told my parents to bring me here because he also had suffered a mental issue 13 years ago, he was treated from Butabika hospital and he was proud of it.”* ***male participant***
>
> *“My father’s friend knew I have got a problem with my head, he told them and convinced them that she doesn’t need to be taken to the traditional healers or to the church and she is prayed for any more but we have to take her to Butabika because that’s where we can get the doctor and proper treatment that is why they brought me here. I did not need to be taken back to the traditional doctor or to the church but they had to bring me here in Butabika”*. ***female participant***.
>
> *“I am used of taking people to hospital because I am a policer officer and I always take those people. So, my option when it comes to the persons who is closer to me and they have issues similar to those we always bring here, that was the only option I had left because we had tried all the other things”* ***male caregiver***.

#### b. Fear of stigma associated with illness

In other cases where ACT was not effective, participants had to move from one service provider to another to pursue a lasting solution—however, this movement from one service provider to another triggered stigma in the community.

> *“When everything [ACT] failed, I realized that am going to be a laughingstock in the community because my relative had started to move around naked in the community, and I knew that it will make me feel bad. What I did I went to the LC (*LC is a short form of local council which is the smallest administrative unit in Uganda) and they wrote for me a letter and I went to the police. The police told me to pay their fuel to take her to Butabika hospital.”* ***male caregiver***.
>
> *“…because in the community where we stay, everyone speculates. Some say that it is witchcraft others it is spiritual. One suggests a witch doctor another suggests a different traditional healer. You try many options so you end up in many places and feel you are stigmatized, even in the church they pray for you when everyone is seeing you so you feel ashamed. Butabika was a better option and you are assured because it’s where the experts are and people there are in similar circumstances.”* ***female caregiver***

## Discussion

Our main findings were that the reasons for a switch from alternative and complementary therapy to evidence-based treatments were similar among FEP patients and their caregivers apart from societal stigma, which was a key reason for switching in only caregivers.

### Catastrophic expenditures as a reason for the switch to evidence-based treatments

Patients with psychotic disorders switched to EBTs due to the cost of ACT. There were no free “public” ACT therapies like at government hospitals. Patients were willing to pay for services if there were positive results and a transparent payment structure. As government financing is often limited for mental health services in low resource settings (25-27), the acceptability of cost-sharing needs review as families may accept payment as long as there are good outcomes (28). As affordable and accessible treatment contributes to proper care and timely resolution of symptoms, the need for mental health insurance may be helpful. Uganda has no government health insurance policy, and private insurance policies sometimes exclude mental health services (29). Since a national insurance health policy is soon to be rolled out, mental health services must not be excluded as patients may not be able or willing to pay for long term care. The role of community-based health insurance needs review (29, 30).

### Poor treatment outcomes as a reason for a switch

Patients reported insufficient improvement of the illness while using ACT. This is in keeping with previous literature highlighting the inadequate response to treatment in patients treated with ACT in Uganda (1, 31). It is recommended that patients with psychotic disorders be treated with antipsychotic medication early in the illness at the first episode of psychosis (21, 32, 33). These delays in initiating care are drivers for long durations of untreated psychosis and corresponding poor long term outcomes. Studies are greatly needed on interventions to reduce the DUP in patients using ACT.

### Societal support and stigma driving change to Western medicine

In many cases, the recommendation to switch to EBTs were provided after ACT had been tried and failed. ACT for psychosis treatment is less stigmatized than EBTs, as described in previous literature in the same setting (2, 6). In many cases, the community was a crucial driver in referrals to Butabika hospital for EBTs. Friends, family, former patients with lived experiences and non-mental health workers were drivers of switching to evidence-based treatments. Caregivers only accepted the need to try evidence-based treatments to avoid the stigma associated with failure to recover with ACT. It may explain why although many patients in this setting used ACT, only the severe patients are reviewed in hospitals (19, 34). At a population level, interventions destigmatizing EBTs for psychotic disorders may be accepted by communities. There is a need to educate the public that evidence-based treatments do not need to be after failed attempts with ACT. Community acceptance and support of evidence-based treatments is a crucial opportunity to reducing DUP.

### Need to understand the illness as a reason for a switch

It has been suggested that ACT is much better at explaining cultural idioms of distress (2). We did not find evidence to support this theory and found that patients and caregivers switched to evidence-based treatments after ACT failed to explain the causes of the illness. There is a need to simplify the theories surrounding psychosis and include it as part of psychosocial management for psychotic disorders in this setting.

### Limitations

The study cannot explain why some participants use ACT only and do not switch to evidence-based treatments or if, after using evidence-based treatments, patients switch back to ACT. These are all critical findings to reduce DUP. As our sample was purposefully selected to include patients who had switched, further studies are required.

### Conclusion

The study showed that for many patients and caregivers, ACT is the initial treatment of choice for patients with psychotic disorders and their caregivers even in the presence of evidence-based treatment. Further studies on collaboration between evidenced-based treatment providers and ACT providers are recommended to ensure earlier initiation of antipsychotic medication which has vast implications for reducing the duration of untreated psychosis and patient outcomes for psychotic disorders. As many patients and caregivers may opt for ACT as their first treatment choice, it is also essential to better understand ACT to further these collaborations in the management of psychotic disorders This could be in the form of building better networks with ACT providers or joint education or awareness programs (2). There is a need for even greater access to mental health treatment to address delay in treatment for FEP. Provision of accessible good quality atypical antipsychotic medication with fewer side effects is recommended as patients are eager to use cheap and effective services.

## Data Availability

The codebook analyzed during the current study is available from the corresponding author

## Ethical considerations

These have been previously described (22). Briefly, ethical approval was received from ethics committees at the school of medicine research and ethics committee of the college of health sciences at Makerere University (#REC REF 2017–153) as well as the Uganda National Council of Science and Technology (HS237ES). Informed consent was obtained from all study participants before enrollment and signed consent forms are available. Participants and caregivers were each reimbursed $6 (approximately sh20 000) for their time, either on completion of the full study assessment or when consent was withdrawn. No individual participant data has been reported.

## Author contributions

EKM conceptualised the research idea. EKM, RL, NN, LO, AS and SM were responsible for determining the content and scope of the study. EKM, LO, AS and SM were responsible for determining the design and methods of the study. NN and RL were responsible for determining the study tools and were content experts. ASS and AJN performed the in-depth interviews and data analysis. All authors were also involved in writing this protocol and accepted the final version.

## Data availability

The codebook analyzed during the current study is available from the corresponding author.

